# Uromonitor®: Clinical Validation and Performance Assessment of a Urinary Biomarker for Recurrence Surveillance in Non-Muscle Invasive Bladder Cancer Patients

**DOI:** 10.1101/2023.11.02.23297958

**Authors:** Pedro Ramos, João Paulo Brás, Carolina Dias, Mafalda Bessa-Gonçalves, Hugo Prazeres, Francisco Botelho, João Silva, Carlos Silva, Luís Pacheco-Figueiredo

**Affiliations:** Departamento de Urologia, Centro Hospitalar e Universitário São João, Porto, Portugal; Instituto de Investigação em Ciências da Vida e Saúde - ICVS/3bs Laboratório Associado, Escola de Medicina, Universidade do Minho, Braga, Portugal; Instituto de Investigação e Inovação em Saúde (i3S), Universidade do Porto, 4200-135 Porto, Portugal; Instituto de Patologia e Imunologia Molecular, Universidade do Porto (Ipatimup), 4200-135 Porto, Portugal; Departamento de Cirurgia e Fisiologia, Faculdade de Medicina, Universidade do Porto, Porto, Portugal; Departamento de Urologia, Hospital Privado Braga Sul – Grupo Trofa Saúde, Braga, Portugal

**Keywords:** urinary biomarkers, bladder cancer, cystoscopy, cytology, follow-up, non-muscle-invasive bladder cancer, recurrence, progression

## Abstract

**Introduction:** Bladder cancer (BC) remains the most common malignancy of the urinary tract, with non-muscle invasive BC (NMIBC) representing the vast majority of bladder cancer patients. The current standard of care (SOC) follow-up in NMIBC patients demands an intensive schedule and requires costly and burdensome methods, driving the development of alternative, non-invasive, cost-effective methods that may complement or serve as substitutes to cystoscopy and cytology. Uromonitor® is a urine biomarker test that detects hotspot mutations in three genes (*TERT, FGFR3*, and *KRAS*) for the evaluation of disease recurrence. The aim of the current study was to assess its performance comparing it to the current SOC methods.

**Materials and Methods:** A total of 528 NMIBC surveillances from 439 individual patients were enrolled from December 2021 to June 2023. All subjects underwent SOC methods and provided an urine sample before undergoing cystoscopy for Uromonitor® analysis. Sensitivity, specificity, positive predictive value (PPV) and negative predictive value (NPV) were calculated for recurrence and compared to the gold-standard cystoscopy plus trans-urethral resection (TURBT) pathology.

**Results:** Uromonitor® displayed a sensitivity of 87.2%, with only 6 recurrences failing to be detected by the urinary biomarker test, a specificity of 99.2%, a positive predictive value (PPV) of 93.2% and a negative predictive value (NPV) of 98.8%. Cystoscopy showed a total of 22 (31,88%) false positives not confirmed by TURBT, while Uromonitor® presented only 3 positive tests where no suspected lesions were found in cystoscopy. Sensitivity, specificity and NPV values for Uromonitor® also remained high across all NMIBC grades and stages.

**Conclusion:** In the present study, we confirmed that the Uromonitor® biomarker test represents a reliable tool in the detection of NMIBC recurrence in patients undergoing routine surveillance, regardless of stage and grade. It offers either an alternative or a complement to the current SOC methods, providing rapid results and a non-invasive option, potentially improving patients’ quality of life and helping reduce the economic burden of NMIBC follow-up. To our knowledge, this is the largest single-center study assessing Uromonitor®’s performance and thus validating its usefulness in clinical practice.

## 1. INTRODUCTION

Urinary bladder cancer (BC) is the second most common genitourinary malignancy and stands as the 10th most common worldwide, with an estimate of more than 570,000 new cases and 210,000 deaths in 2020, being the urothelial carcinoma the most frequent histology (1). The risk of BC increases with age, with age-specific curves steeply increasing after the age of 50 years (2). Consequently, the incidence predictions report an expected increased number of new cases in the next decades due to the observed aging of the world population (3).

Since approximately 75% of the patients present with a non-muscle-invasive bladder cancer (NMIBC), which has a favorable prognosis, there is an rising number of prevalent cases. The high rates of NMIBC recurrence and lifetime risk of progression make intensive follow-up mandatory (4,5). The current follow-up schedule of patients with a NMIBC diagnosis includes regular surveillance with cystoscopy combined with urine cytology, as well as upper-tract evaluation with imaging. This follow-up regimen should be maintained years following diagnosis and may even continue throughout life (6). Therefore, there is a rising healthcare burden associated with BC management, with some reports highlighting the highest lifetime costs per bladder cancer patient, among all malignancies (7–9)

Cystoscopy is considered the gold standard to follow-up NMIBC patients since it remains as the surveillance examination with higher levels of sensitivity. However, it has some drawbacks: a) a poorer sensitivity to identify flat lesions, carcinoma in situ (CIS) and microscopic lesions; b) it is an invasive and uncomfortable procedure, inducing anxiety and carrying a risk of infection; c) it is a time-consuming procedure posing a relevant burden in healthcare resources (10). On other hand, the urinary cytology, although less expensive and noninvasive, it provides substantially lower levels of sensitivity (ranges from 16% in low-grade patients up to 84% in patients in high-grade disease), posing some limitations as using it as a sole method for surveillance of NMIBC patients. The latter is the reason why this exam is only recommended as a complement to cystoscopy in patients with high-grade disease (11).

Therefore, there is a growing interest in BC research for a new, more cost-effective, less invasive, and accurate follow-up method to use in NMIBC patients. The urinary biomarkers surge as promising tool to be used in NMIBC surveillance programs, both decreasing the need of cystoscopy withing some follow-up time frames and replacing the cytology as more accurate exam to be used in complement with the cystoscopy. Several urinary biomarkers have been developed and received FDA approval through the years. However, tests like the NMP22 BladderCheck, UroVysion, or BTA stat have not been successfully implemented into clinical practice due to insufficient NPV and low specificity (12).

More recently, new urinary biomarkers have demonstrated a higher NPV in small samples of NMIBC cases (13). Among them, Uromonitor® emerged as one with an expected better diagnostic accuracy. This test evaluates a subset of hotspot alterations in three different types of genes (TERT, FGFR3 and KRAS), that are most associated with NMIBC (14).

Encouraging results have been obtained with Uromonitor®. In the first study, Uromonitor® showed a sensitivity of 73.5% and a specificity of 73.2% in detecting TURBT confirmed recurrences, analysed among 331 urine samples collected at 18 different centers. With the addition on KRAS hotspot mutation analysis, sensitivity increased to 100% in the detection of TURBT-confirmed recurrence, with a specificity of 83.3% (15). Later, Sieverink *et al*. performed the first clinical validation study with Uromonitor® incorporating TERT/FGFR3/KRAS full panel. A total of 97 patients with history of NMIBC were included. Uromonitor® presented an overall sensitivity of 93.1%, a specificity of 86.8%, a positive predictive value (PPV) of 75.0%, and an NPV of 96.7% (16). In a recent clinical trial focusing on low grade NMIBC, including 171 patients and 380 cystoscopies/Uromonitor® tests, Uromonitor® demonstrated a sensitivity of 89.7%, specificity of 96.2%, a negative predictive value (NPV) of 98.8%(17). Despite these promising results, further evidence and clinical practice validation are still needed for Uromonitor® to establish itself as a reliable alternative tool for early recurrence detection during NMIBC follow-up. In this study, we aimed to validate these results in a larger prospective independent cohort of NMIBC patients undergoing follow-up in a high-volume BC center.

## 2. MATHERIALS AND METHODS

### 2.1 Study Design

A prospective, observational, single-center study was implemented. We included patients with a previously diagnosed NMIBC under follow-up at São João University Hospital Center in Porto, Portugal. All selected patients in the study collected urine samples for Uromonitor® testing before undergoing flexible cystoscopy at the hospital’s outpatient clinic. Patients were enrolled from December 2021 to June 2023.

Inclusion criteria were the following: ≥18 years of age; history of NMIBC currently under follow-up; being able to give written consent; being able to provide a minimum of 10mL of urine prior to undergoing standard-of-care (SOC) cystoscopy.

Exclusion criteria: undergoing cystoscopy for a different diagnosis than BC; previous diagnosis of muscle-invasive bladder cancer (MIBC); previous diagnosis of upper tract urothelial carcinoma (UTUC); providing inadequate material for Uromonitor® testing.

### 2.2 Patient Data Collection

Clinical information was collected from all patients enrolled, including: age, gender, smoking history (ever smoker/never smoker), date of NMIBC diagnosis (1^st^ TURBT), symptoms at presentation (hematuria/other symptoms/radiologic incidentaloma via ultrasound or tomography), size of primary BC lesions (<1cm/ 1-3cm/ >3cm), type of lesion (polypoid/sessile/flat), stage at 1^st^ TURBT (pTa/ pT1) plus 2nd look TURBT if appliable, grade of primary lesion (HG/ LG), histology (urothelial carcinoma (UC)/ variant histology (VH)) and adjuvant intravesical therapy (chemotherapy or BCG). The cystoscopy and subsequent transurethral bladder tumor resection (TURBT) results (when applied) were also collected. The Uromonitor® dichotomic results (positive/negative) were registered as well as the individual hotspot mutations identified in each case (TERT - 124/ TERT -146/ FGFR3 248/ FGFR3 249/ KRAS 12/13 / KRAS 61).

All urine collections for Uromonitor® assessment were performed within the standard clinical NMIBC follow-up scheme in which all patients came in for a regular follow-up cystoscopy. Every patient was asked to provide additional urine for study purposes, immediately before cystoscopic evaluation. Following urine collection, all patients underwent a standard of care cystoscopy. Recurrence or progression were defined as those in which the cystoscopy revealed suspicious lesions and were further pathologically proven after TURBT.

### 2.3 Urine Collection, Sample Processing and Testing

Specimens were processed using the Uromonitor® IVD Test (U-Monitor, Porto, Portugal). Uromonitor® is composed by three subunits. The “Uromonitor® – Urine filtering kit” (#Urokit1) provides the components needed for sample collection. The principle of the procedure relies on the filtration of urine samples. Using a syringe, urine samples pass through a filter, with a proprietary conservative, where exfoliated bladder cells are trapped and enriched for subsequent procedures. The “Uromonitor® - DNA Extraction and preparation kit” (#Urokit2) provides the reagents for the extraction and purification of genomic DNA. The “Uromonitor® - Real-Time PCR kit for the amplification and detection of TERT, FGFR3 and KRAS hotspot mutations” (#Urokit3) provides an efficient and fast method for the amplification and detection of mutations in the TERT promoter (-124 and -146), FGFR3 codons 248 and 249, and KRAS codons 12/13 and 61, through Real-Time PCR.

#### 2.3.1 Urine filtration

Urine samples collected from patients prior to flexible cystoscopy were filtered using #Urokit1, following their consent. Samples of 10–20 mL were filtered within an hour of collection and stored at 4 °C. Filters containing the samples were sent on a weekly basis to U-Monitor Lab (UPTEC, Porto) for analysis. The company was blinded to the flexible cystoscopy results and received samples with ID numbers, which were stored in a secure database in the Department of Urology in São João University Hospital Centre.

#### 2.3.2 DNA Extraction

Filtered urine samples underwent final analysis in U-Monitor Lab (UPTEC Asprela, Porto). DNA was extracted using #Urokit2 and prepared for Real-Time PCR. Briefly, filters were inverted and placed on top of the cell lysis tubes. Syringes with 400μL cell lysis buffer were attached to the filter, and the buffer filtered into the cell lysis tube. Cell lysis tubes were then incubated at 60 °C for 30 min under agitation. After this, proteinase K was added, and the mixtures incubated for another 10 min at 60 °C. Then, 450 μL of normalizing solution were added and vortexed for 5 seconds. The lysate mixtures were transferred to the respective spin columns and centrifuged for 1 min at 3300g. After that, 2-step washing centrifugations were performed. Finally, spin columns were transferred to the respective elution tubes, and 50 μL of elution buffer were added. After a 5-minute incubation at RT, tubes were centrifuged, and the eluates containing the DNA stored at -20°C until Real-time PCR analysis.

#### 2.3.3 Real-time PCR

The extracted DNA was amplified and detected on a qPCR real-time machine (StepOnePlus™, Thermo Fisher Scientific, Waltham, MA, USA) using the proprietary chemistry for amplification and detection, as provided with the #Urokit3. It contains 3 independent assays that use allele-specific primers in a multiplex reaction to identify the presence of TERTp, FGFR3 and KRAS mutations in a total of 6 reactions per sample. Each reaction contains primer sets and probes for detection of the mutations, as well as the endogenous control genes. The list of mutations detected by the Uromonitor is provided in Supplementary Table 1. Multicomponent and amplification signals were analysed using StepOne™ Software 2.3 as recommended by the manufacturer (U-Monitor, Porto, Portugal). If at least one of the screened alterations provided a positive result, then the test was positive.

### 2.4 Statistical analyses

The assessment of the Uromonitor® accuracy was performed estimating the sensitivity, specificity, and positive and negative prediction values. For assessment of differences between groups, the Fisher exact test, Student's t-test, or Mann-Whitney test, were applied according to variables and groups. A p-value <0.05 was considered as statistically significant, and the Confidence Interval (CI) used was 95%.

### 2.5 Ethics statement

This study was an observational study, with no interference from the investigators in the standard of care follow-up procedures. The subjects did not need to have extra visits to the hospital, since the urine collection was performed previously to the cystoscopy. Patients were invited to participate in the study, and they were included only after signing an informed consent form. The patients’ data was stored after anonymisation, and the encryption key remained safely stored in the hospital. All procedures described in this study were in accordance with national and institutional ethical standards and the Declaration of Helsinki. Procedures were previously approved by Ethical Review Committee in University Hospital Centre of São João.

## 3. RESULTS

A total of 553 urine samples were collected from 464 individual patients throughout the study’s timeline, which ranged from December 2021 to May 2023. Twenty-five samples were excluded: 19 were collected from patients without previous BC/UTUC history and 6 were from patients undergoing cystoscopy during UTUC follow-up. Therefore, 528 samples from 439 patients were deemed eligible for the study. Out of the 528 SOC cystoscopies performed on eligible cases, 69 (13.1%) exhibited suspected recurrence. This suspicion was confirmed in 47 cases by TURBT, and within this group, 2 patients showing progression to MIBC according to the TURBT’s pathology report. Among the 47 pathologically proven recurrence cases, 31 (66.0%) presented pTa lesions, 14 (29.8%) had pT1 tumors, and 2 (4.3%) patients progressed to pT2+ disease. Further stratified by both stage and grade, 4 (8.5 %) had pTa low-grade, 27 (57.5%) had pTa high-grade, 14 (29.8%) had pT1 high-grade, and 2 (4.3%) progressed to pT2. Carcinoma in situ was not reported in any of the recurrent histologies (Table 1).

**Table 1.** Descriptive data of the cohort of 439 NMIBC cases.

| Variables | N=439 patients |
| --- | --- |
| Age (years)* | 71.0 (63.0-77.0) |
| Follow-up time (months)* | 29.8 (12.8-55.0) |
| Gender<sup>‡</sup> | Male: 368 (83.8)<br>Female: 71 (16.2) |
| Smoking status<sup>‡</sup> | Ever smoker: 319 (72.7)<br>Never smoker: 120 (27.3) |
| Symptoms at presentation<sup>‡</sup> | Hematuria: 354 (80.6)<br>Other symptoms: 46 (10.5)<br>Asymptomatic: 39 (8.9) |
| Type of lesion<sup>‡</sup> | Polypoid: 369 (84.3)<br>Sessile: 51 (11.6)<br>Flat: 18 (4.1) |
| Size of lesion<sup>‡</sup> | < 1cm: 126 (28.7)<br>1 – 3cm: 208 (47.4)<br>> 3cm: 105 (23.9) |
| Stage at diagnosis<sup>‡</sup> | pTa: 262 (59.7)<br>pT1: 172 (39.2)<br>CIS: 5 (1.1) |
| Grade at diagnosis<sup>‡</sup> | Low grade: 124 (28.3)<br>High grade: 315 (71.8) |
| Histology<sup>‡</sup> | Urothelial carcinoma: 431 (98.2)<br>Variant histology: 8 (1.8) |

\* median (Percentile 25-Percentile 75)
<sup>‡</sup> n (%)

Out of the 528 eligible samples tested, 44 (8.3%) yielded a positive result on Uromonitor® (UM+) for one or more hotspot mutations, while the remaining 484 (91.7 %) produced negative results. Among the 44 patients in the UM+ group, 41 (93.2%) were confirmed to have a pathology proven recurrence. However, the remaining 3 UM+ patients had a previous history of NMIBC, but did not undergo TURBT because SOC follow-up methods did not raise suspicion. All three patients were followed-up for more than 12 months without any recurrence suspicion detected thus far.

In terms of Uromonitor®'s overall performance it exhibited a sensitivity of 87.3%, with 6 recurrences remaining undetected, a specificity of 99.3%, a positive predictive value (PPV) of 93.2% and a NPV of 98,8% (Table 2).

**Table 2.** Performance of different tools in detection of recurrence in NMIBC follow-up in all stages/grades.

| Stage and grade at diagnosis |  | Uromonitor® |
| --- | --- | --- |
| pTa Low grade | Sensitivity | 75.0% |
|  | Specificity | 100.0% |
|  | PPV | 100.0% |
|  | NPV | 99.2% |
| pTa High grade | Sensitivity | 87.5% |
|  | Specificity | 98.7% |
|  | PPV | 87.5% |
|  | NPV | 98.7% |
| pT1 High grade | Sensitivity | 75.0% |
|  | Specificity | 100.0% |
|  | PPV | 100.0% |
|  | NPV | 99.2% |
| ALL<br>(Including CIS) | Sensitivity | 87.2% |
|  | Specificity | 99.2% |
|  | PPV | 93.2% |
|  | NPV | 98.8% |

We conducted a subset analysis to evaluate the performance of Uromonitor® across different stages and grades of NMIBC (Table 2): pTa LG – sensitivity 75.0%, specificity 100.0%, PPV 100%, NPV 99.2%; pTa HG sensitivity 87.5%, specificity 98.7%, PPV 87.5%, NPV 98.7%; pT1 HG - sensitivity 75.0% specificity 100.0% PPV 100.0% NPV 99.2%. Additionally, in a subset of patients with suspected recurrence in cystoscopy, Uromonitor® demonstrated an overall sensitivity of 87.3%, specificity of 100.0%, PPV of 100.0% and NPV of 78.6%.

Furthermore, cystoscopy revealed a total of 22 (31.9%) false positives. These patients with suspected recurrence underwent TURBT, where no malignancy was found after pathology analysis.

## 4. DISCUSSION

The challenging nature of NMIBC follow-up can be attributed to both its high recurrence rates as well as the risk of progression to MIBC. Conversely, the observed progression rates in these patients can, in part, be attributed to limitations of current follow-up methods. Urinary cytology, while offering high specificity for high-grade disease and a non-invasive approach with minimal associated morbidity, falls short in terms of sensitivity, especially when detecting low-grade UC, resulting in a high false-negative rate (11). Additionally, its interpretation is subjective and can introduce significant interobserver variability, compromising its reliability (18). Moreover, cytology cannot distinguish between benign inflammatory changes and malignancy, leading to false-positive results and unnecessary invasive procedures (19). Lastly, the accuracy of cytology can be influenced by urine collection and processing techniques, making it susceptible to sampling errors (20).

Cystoscopy remains a highly sensitive and reliable method for early recurrence detection in NMIBC. It enables direct visualization and accurate assessment of tumour size, location, multifocality, and appearance, while also offering the option for immediate biopsy for histologic characterization. However, it is an invasive procedure that can cause patient discomfort and morbidity. Furthermore, it relies on specialized training and equipment, which can make it costly and less accessible in certain healthcare settings(21).

Considering all the information discussed above, it becomes evident why there is a growing interest in alternative, cost-effective, non-invasive methods with high sensitivity and specificity for follow-up surveillance in NMIBC patients. These tools can either serve as alternatives to or complement the current follow-up schedule, potentially reducing the need for frequent cystoscopies and replacing urinary cytology. Urinary biomarkers appear to be a promising alternative in this context due to their simplicity, non-invasiveness, and cost-effectiveness. However, currently approved biomarkers have failed to find widespread clinical implementation due to issues such as low specificity and inadequate negative predictive values (NPV), leading to a high rate of false-positive cases attributed to the design of their arrays (12). Uromonitor®, among other novel urinary biomarker tests (UBT’s), has emerged displaying higher accuracy (mainly specificity and NPV) in the NMIBC surveillance (15,16). These advancements offer a promising and safe tool to alleviate the burden of frequent cystoscopies across all risk groups of NMIBC patients (22).

Within our study, we observed that Uromonitor® had high levels of overall sensitivity (87.2%) and NPV (98.8%), demonstrating its reliability as a surveillance tool with a small number of false negatives (n=6). To further elucidate the reasons behind those misses we collected the Formalin-Fixed Paraffin-Embedded (FFPE) tissue from the original TURBT of those six cases and analyzed the presence of TERTp, FGFR3 and KRAS mutations with “Uromonitor® - Real-Time PCR kit for the amplification and detection of TERT, FGFR3 and KRAS hotspot mutations”. Analyses revealed that 6 out of these 6 samples presented at least one of the mutations analyzed within the Uromonitor®. Interestingly, this excludes the hypothesis of these cases being missed at first, because they bear mutations not detected by the Uromonitor®. Instead, these results rather indicate that: (i) there might have been a problem with sample filtration (i.e. <10 mL filtered), that turned out to provide insufficient detectable mutated material; (ii) or that for these samples, the amount of mutated material present in the 10 mL of urine filtered, was below the detection limit of Uromonitor®.

Uromonitor® also demonstrated high levels of specificity (99.2%) and PPV (93.2%), demonstrating the usefulness of this instrument to confirm the diagnosis of recurrence or progression. In fact, we observed that amidst the 69 cases with suspected recurrence in cystoscopy, 22 revealed no evidence of malignancy on TURBT pathology analysis. Among these patients, Uromonitor® yielded negative results in all 22 cases. Most of those false-negative cystoscopic results were associated with equivocal findings associated with the inflammation of adjuvant intravesical treatments (bacillus Calmette-Guérin (BCG) or chemotherapy). This suggests that if Uromonitor® had been used as an adjuvant tool to confirm suspicious lesions observed in cystoscopy evaluation, approximately 30% of TURBT’s could had been avoided, mitigating the associated morbidity and costs.

Among the 44 patients with positive Uromonitor® results, three (6.8%) showed negative cystoscopy evaluations. These results may constitute false positives, but one cannot exclude the possibility that these patients had either a recurrence too small to be observed with cystoscopy or a recurrence in the upper urinary tract. After more than 6 months of surveillance, none of these patients had presented signs of the disease as assessed by the SOC protocol, which increases the likelihood of constituting false positives. A longer follow-up is required to fully understand the false positive rate of Uromonitor® for late recurrences.

The distribution of hotspot mutations identified among the 44 positive Uromonitor® tests is displayed in Table 3. The prevalence of hotspot mutations detected was according to previous data described in NMIBC (23–25). Moreover, evidence speculating the role of hotspot mutations as a prognostic marker in NMIBC has accumulated over the years, particularly for FGFR-3, regarding both risk of recurrence/progression and response to adjuvant intravesical therapy, such as BCG or immunotherapy (26–28). Again, FGFR3 assumes an important role as a predictive biomarker due to the development of FGFR3-targeted therapies - the use of FGFR3 signaling blockage using FGFR inhibitors, such as erdatifinib and infigratinib has have shown promising results in clinical trials for advanced NMIBC with FGFR3 alterations (29,30). Erdatifinib, a FGFR1-4 inhibitor, has already been granted FDA approval, and is in use in patients with metastatic BC (31).

**Table 3.** Distribution of hotspot mutations identified by Uromonitor®.

| **Hotspot mutation detected by Uromonitor®** | **Total number of cases detected (%)** |
| --- | --- |
| TERT 124 | 16 (36.3%) |
| TERT 146 | 6 (13.6%) |
| FGFR3 - 248 | 19 (43.2%) |
| FGFR3 - 249 | 3 (6.8%) |
| KRAS 12x/13x | 3 (6.8%) |
| KRAS 61x | 2 (4.5%) |
| **Combinations of hotspot mutations detected by Uromonitor®** | **Total number of cases detected (%)** |
| TERT 124 + FGFR3-249 | 1 (2.27%) |
| TERT 124 + FGFR3-249 + KRAS 61x | 1 (2.27%) |
| TERT 124 + FGFR3-248 | 1 (2.27%) |
| TERT 124 + TERT 146 | 1 (2.27%) |

Summing the previous discussed main findings of this study, Uromonitor® show the potential for widespread implementation in clinical practice in the near future, although cystoscopy cannot be entirely abandoned and will continue to play an essential role during the follow-up of NMIBC. In high-risk cases, remains highly unlikely that practitioners would rely solely on biomarkers to assess recurrence and progression. For these patients, Uromonitor® could be used not only as a complement to current methods but also as a potentially risk stratifying tool to assess tumor aggressiveness and guide treatment intensification and planning. Studies evaluating the correlation between hotspot mutations and disease aggressiveness are still scarce, yet some evidence suggests that these genetic aberrations can impact disease behavior in BC (4,32,33).

In low- and intermediate-risk NMIBC patients, Uromonitor® could significantly alleviate the burdensome impact of follow-up by reducing patient discomfort, morbidity, and substantial costs associated with cystoscopy. This could be achieved by implementing an alternating approach between cystoscopy and Uromonitor® testing, thus reducing the overall frequency of cystoscopies without compromising patient’s safety.

Additionally, employing this test when dubious or equivocal lesions are observed during cystoscopy - an occurrence that is fairly common in clinical practice and can induce anxiety in both physicians and patients - could aid in deciding whether a patient should undergo invasive procedures such as TURBT.

Another advantage provided by this test is its reproducibility, as it relies on qPCR, a methodology that is well-established in most laboratories. This enables in-house testing using readily available technology, user-friendly components, and affordable equipment, without the need for specialized technicians. The test delivers same-day results within a short timeframe (six hours) and provides a clear binary positive-negative result.

Despite the promising results, the authors acknowledge the following limitations of the study: a) the relatively low number of histological confirmed recurrences compared to the performed cystoscopies may have limited the study statistical power; b) despite encompassing over 500 NMIBC follow-up outpatient visits, which included cystoscopy and Uromonitor® testing for 439 individual patients, the overall sample remained limited to a single-center, with its own clinical practices and treatment protocols (although in accordance with international guidelines), limiting the potential for extrapolating the results to other healthcare settings, with different patient populations and practices. On other hand, this study, as far as the authors know, has one of the largest prospective cohorts of NMIBC patients assessed with a UBT and the largest one using Uromonitor®.

Further research, involving a larger cohort of NMIBC patients through a multicentre trial comparing Uromonitor® against the SOC, should be conducted to definitely determine whether Uromonitor® could serve as a safe replacement of the current follow-up protocol. Additionally, there is a need for new evidence regarding quality-of-life analyses and the cost effectiveness of employing this biomarker. Nevertheless, the current study adds clinically relevant knowledge towards validating and subsequentially integrating Uromonitor® into daily clinical practice.

## 5. CONCLUSION

In the present prospective cohort study, we have confirmed that the Uromonitor® biomarker test, with its high accuracy, represents a reliable tool in the detection of NMIBC recurrence/progression in patients undergoing routine surveillance, regardless of stage and grade. It offers both an alternative or a complement to the current SOC methods, as it provides rapid results and a non-invasive option, potentially improving patients’ quality of life by reducing the need for uncomfortable cystoscopies. An analysis of cost-effectiveness appears to be the next logical step for wide clinical practice integration.

**Figure 1.**
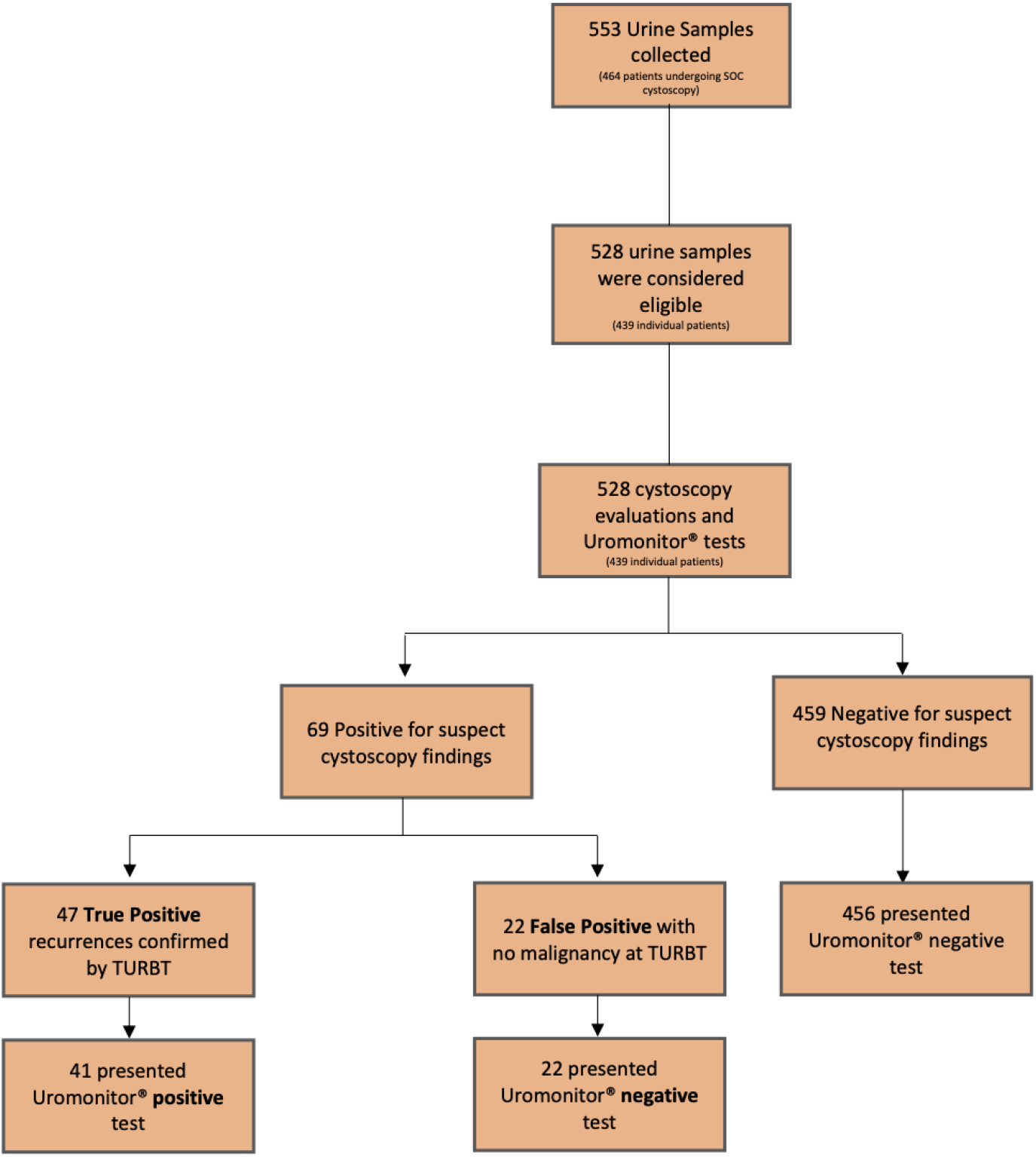
A flowchart displaying the Uromonitor® study’s findings

## Author Contributions

Methodology: P.Ramos, H.Prazeres, F. Botelho, C.Silva, L. Pacheco-Figueiredo

Data curation: P.Ramos, J. Paulo-Brás

Writing—Original draft: P.Ramos, J. Paulo-Brás, L. Pacheco-Figueiredo

Writing—Review & Editing: P. Ramos; J. Paulo-Brás, C. Dias, M. Bessa-Gonçalves, H. Prazeres, F. Botelho, J. Silva, C. Silva, L. Pacheco-Figueiredo

Supervision: H.Prazeres, L. Pacheco-Figueiredo

All authors have read and agreed to the published version of the manuscript.

## Institutional Review Board Statement

The study protocol was approved by the Ethics Committee

## Informed Consent Statement

Patient consent was waived according to the permission obtained from Ethical Committe.

## Data Availability Statement

The data presented in this study are available in this article.

## Supplementary files

### FFPE tissue processing and analysis

DNA was extracted using GRS Genomic DNA Kit - BroadRange (GK06.0100; GRISP, Portugal) and the presence of TERTp, FGFR3 and KRAS mutations analyzed with the “Uromonitor® - Real-Time PCR kit for the amplification and detection of TERT, FGFR3 and KRAS hotspot mutations” (#Urokit3). Briefly, slides containing 20 um thick tissue slices were deparaffinized using xylene and ethanol. Deparaffinized tissue slices were then microdissected at the microscope into a 1.5 mL tube. Samples were incubated overnight at 60°C with Buffer BR1 and Proteinase K. After that, Buffer BR2 was added and homogenized with the samples. Lysates were then centrifuged for 2 min at 16000g and supernatants mixed with absolute pro-analysis ethanol before being transferred to the “genomic mini spin” columns. Lysate mixtures were centrifuged for 1 min at 16000g. After that, 2-step washing centrifugations were performed. Finally, 25 μL of elution buffer were added, and after a 5-minute incubation at RT, tubes were centrifuged, and the eluates containing the DNA stored at -20°C until Real-time PCR analysis. Real-time PCR analysis was performed as for the urine samples, following #Urokit3 instructions.

